# Persisting antibody response to SARS-CoV-2 in a local Austrian population

**DOI:** 10.1101/2020.11.20.20232140

**Authors:** Dennis Ladage, Delia Rösgen, Clemens Schreiner, Dorothee Ladage, Christoph Adler, Oliver Harzer, Ralf J. Braun

## Abstract

The severe acute respiratory syndrome coronavirus 2 (SARS-CoV-2) has caused a global pandemic. The prevalence and persistence of antibodies following a peak SARS-CoV-2 infection provides insights into the potential for some level of population immunity. In June 2020 we succeeded in testing almost half of the population of an Austrian township with a higher incidence for COVID-19 infections. Now we performed a follow-up study to reassess the prevalence of SARS-CoV-2-specific IgA and IgG antibodies. In 121 people, including 68 participants of the previous study we found the prevalence of IgG and IgA antibodies remaining remarkably stable with 84% of our cohort prevailing SARS-CoV-2-specific antibodies, which is only a slight decrease from 93% four months before. Most patients with confirmed COVID-19 seroconvert, potentially providing immunity to reinfection. Our results suggest a stable antibody response that we observed for at least six months post infection with implications for developing strategies for testing and protecting the population.

---

The world is still challenged by the coronavirus (SARS-CoV-2) pandemic crisis with the second wave culminating in autumn 2020 all over Europe, including Austria. It is still controversially discussed to which extend and for how long previously affected people are immune to a recurring infection. During the cause of an infectious disease, first immunoglobulin M (IgM) and immunoglobulin A (IgA) antibodies are produced by B lymphocytes, which are later replaced by immunoglobulin G (IgG) antibodies. Persisting IgG antibodies are essential for developing a long-lasting immune response. In fact, more than 90% of people with known SARS-CoV-2 infections robustly develop antibodies to the SARS-CoV-2 spike protein, which comprises the receptor binding domain (RBD), enabling the virus to access human target cells (Amanat et al., 2020;Long et al., 2020a;Premkumar et al., 2020;Zhao et al., 2020). Thus, the antibody-based immune response is likely to play a decisive role in immunity to SARS-CoV-2.

In June 2020, we tested almost half of the population of the Austrian township of Weißenkirchen in the Wachau with a reported higher incidence for COVID-19 infections during the first wave in early spring 2020. We used a sensitive laboratory-based ELISA method (Euroimmun), enabling the semi-quantitative measurement of serum levels of IgG and IgA antibodies, specific for the SARS-CoV-2 spike protein. In this pilot study (Ladage et al., 2020), we observed that 12% (99/835) of the tested people were infected and in consequence developed SARS-CoV-2-specific IgG or IgA antibodies (Figure 1A, left panel). 9% (72/835) were positive for IgG antibodies and 9% (75/835) contained IgA antibodies, respectively. In June 2020, 6% (48/835) of our test population were serum-positive for both, SARS-CoV-2-specific IgG, and IgA antibodies.

**Figure 1:**
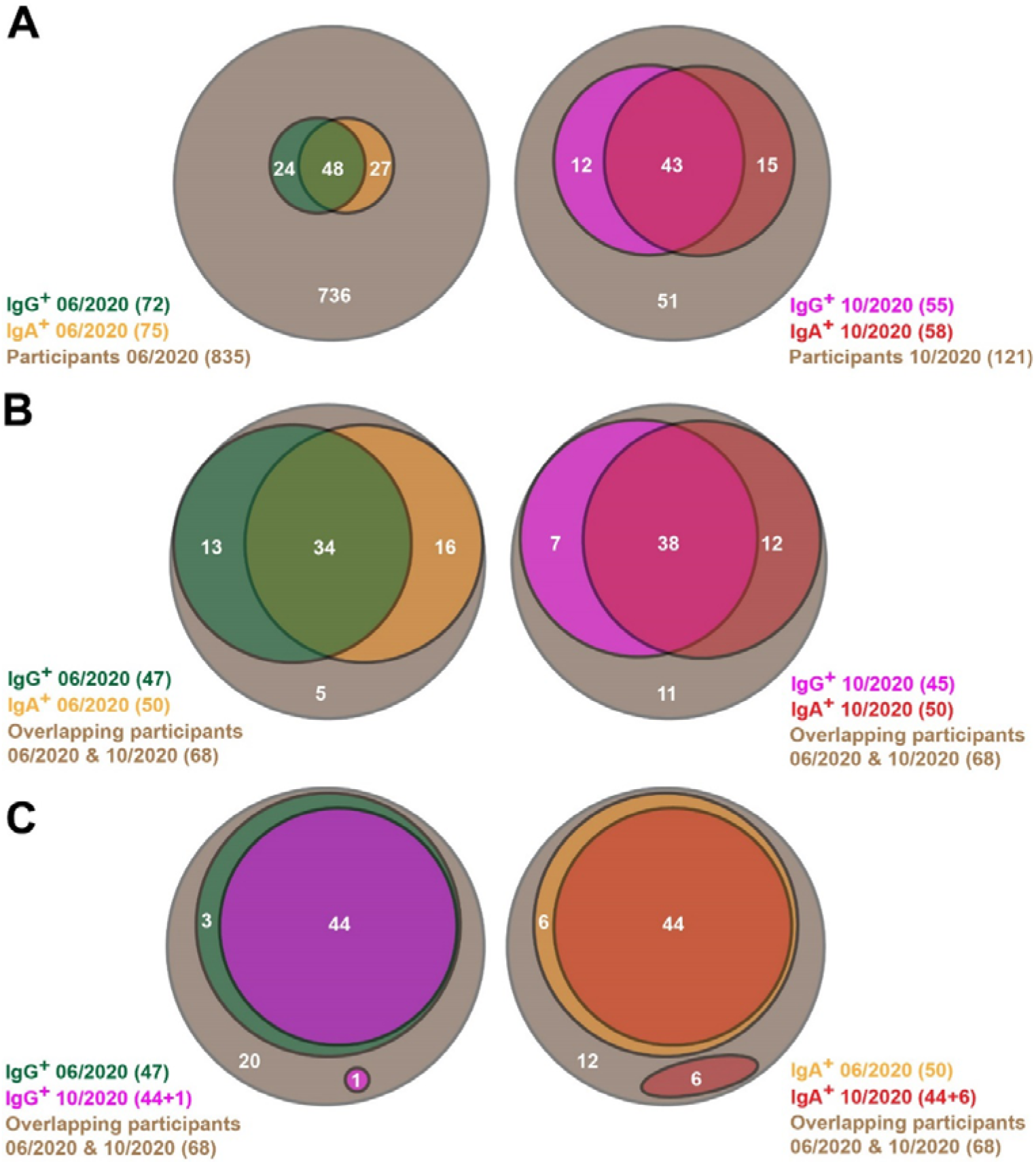
Venn diagrams showing SARS-CoV-2-specific antibody prevalence in the pilot (06/2020) and the follow-up (10/2020) studies. (A) All participants of both studies are shown here. (B, C) Only the participants are considered here, which are involved in both the pilot and follow-up studies.

In October 2020, we performed a small follow-up study to reassess the prevalence of SARS-CoV-2-specific IgA and IgG antibodies in Weißenkirchen and neighboring communities. Blood samples were obtained for detecting SARS-CoV-2-specific IgM and IgG antibodies in an enzyme-linked immunosorbent assay (ELISA) as in the previous study. The study was in accordance with guidelines by the local ethics committee and in approval of local and national authorities. We tested a non-representative group of 121 people, including 68 subjects, which already participated in the pilot study.

Among the 121 participants 58% (70/121) tested positive for SARS-CoV-2-specific IgA or IgG antibodies (Figure 1A, right panel). 45% (55/121) were positive for IgG antibodies and 48% (58/121) contained IgA antibodies, respectively. 36% (43/121) contained both IgG and IgA antibodies. Out of those subjects that already participated 93% (63/68) already tested positive in June 2020 (Figure 1B, left panel). 50% (47/68) were positive for IgG antibodies and 74% (50/68) contained IgA antibodies, respectively. 56% (34/68) contained both IgG and IgA antibodies. In October 2020, we found in 84% (57/68) SARS-CoV-2-specific IgG or IgA antibodies (Figure 1B, right panel). 66% (45/68) contained IgG antibodies and 74% (50/68) contained IgA antibodies, respectively. In 56% (38/68) both classes of antibodies could be found.

Thus, the prevalence of SARS-CoV-2-specific IgG and IgA antibodies remained extremely stable in the re-tested participants (Figure 1B, c.f. left and right panels). After four months we found 84% of our cohort to still have SARS-CoV-2-specific antibodies, which is only a slight decrease from 93% in the previous test in June 2020.

This could be due to a high persistence of individual antibody responses. However, the antibody responses could wane in some individuals, which is superimposed by novel infections in other participants of the same subpopulation. Therefore, we analyzed the changes in antibody prevalence on an individual basis. 94% (44/47) of people with SARS-CoV-2-specific IgG antibodies in June 2020 were still positive for IgG in October 2020 (Figure 1C, left panel). In one person, SARS-CoV-2-specific IgG antibodies could be found the first time in October 2020. 88% (44/50) of participants with SARS-CoV-2-specific IgA antibodies in June 2020 still contained marked IgA levels in October 2020 (Figure 1C, right panel). In six subjects, IgA antibody responses were at first detected in October 2020. Therefore, the persistence of antibody resistance in only marginally influenced by novel infections. When considering the alterations of antibody prevalence on an individual basis, the persistence of antibody responses remained very robust.

Consequently, 97% (33/34) of subjects having both SARS-CoV-2-specific IgG and IgA antibodies in June 2020 still contained significant levels of both classes of antibodies in October 2020 (Figure 2). Notably, only in one subject the IgA antibody levels waned whereas the IgG antibody level remained significantly high. Only three persons with IgG but lacking IgA in June 2020 lost their IgG antibodies by October 2020. Surprisingly, five persons that lack IgA in June 2020 developed IgA by October 2020, then showing both SARS-CoV-2-specific IgG and IgA antibodies. In five persons with IgA but without IgG in June 2020 their IgA antibodies waned by October 2020. Thus, the IgG antibody responses persisted very efficiently from June to October 2020, and the waning of the IgA antibody response was surprisingly low. One would expect a significant loss of the IgA antibodies because they are early and transient responders to an infection followed by the production of the long-lasting IgG antibodies (Isho et al., 2020;Iyer et al., 2020). In contrast, in our study a robust immune response with high levels of both SARS-CoV-2-specific IgG and IgA antibodies guaranteed the most efficient persistence of human antibody response, at least within the first six months after infection.

**Figure 2:**
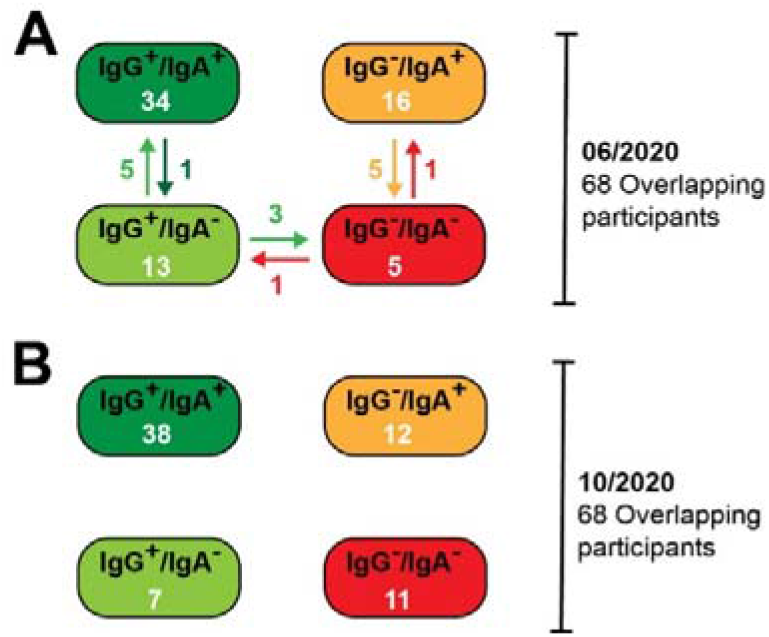
Alterations in the SARS-CoV-2-specific antibody prevalence between the pilot (06/2020) and the follow-up (10/2020) studies. (A) Antibody prevalence in the pilot study. Specific changes are indicated with arrows. (B) Antibody prevalence in the follow-up study.

The SARS-CoV-2-specific serum antibody levels may decrease over time in most individuals, but as long as the signals are above the threshold of the applied ELISA test system, this waning could be missed in our analysis so far. Therefore, we compared the relative IgG and IgA antibody levels from June 2020 and October 2020 for every participant (Figure 3). Using a semi-quantitative ELISA system, both the IgG and IgA antibody levels hardly waned between June and October 2020 (in average 10% for IgG and 14% for IgA). Indeed, in some cases, we even observed increased IgG and IgA antibody levels over time. Thus, these data support our notion that the antibody-based immune responses were very stable in the tested population between June and October 2020. Since most known COVID-19 cases in Weißenkirchen were noted in March 2020, our data propose that the antibody-based immune responses last for more than six months. This may also have implications to the efficiency of SARS-CoV-2 vaccination. A strong antibody-based immune response involving both IgG and IgA antibodies upon vaccination, may be predictive for immunity for more than six months.

**Figure 3:**
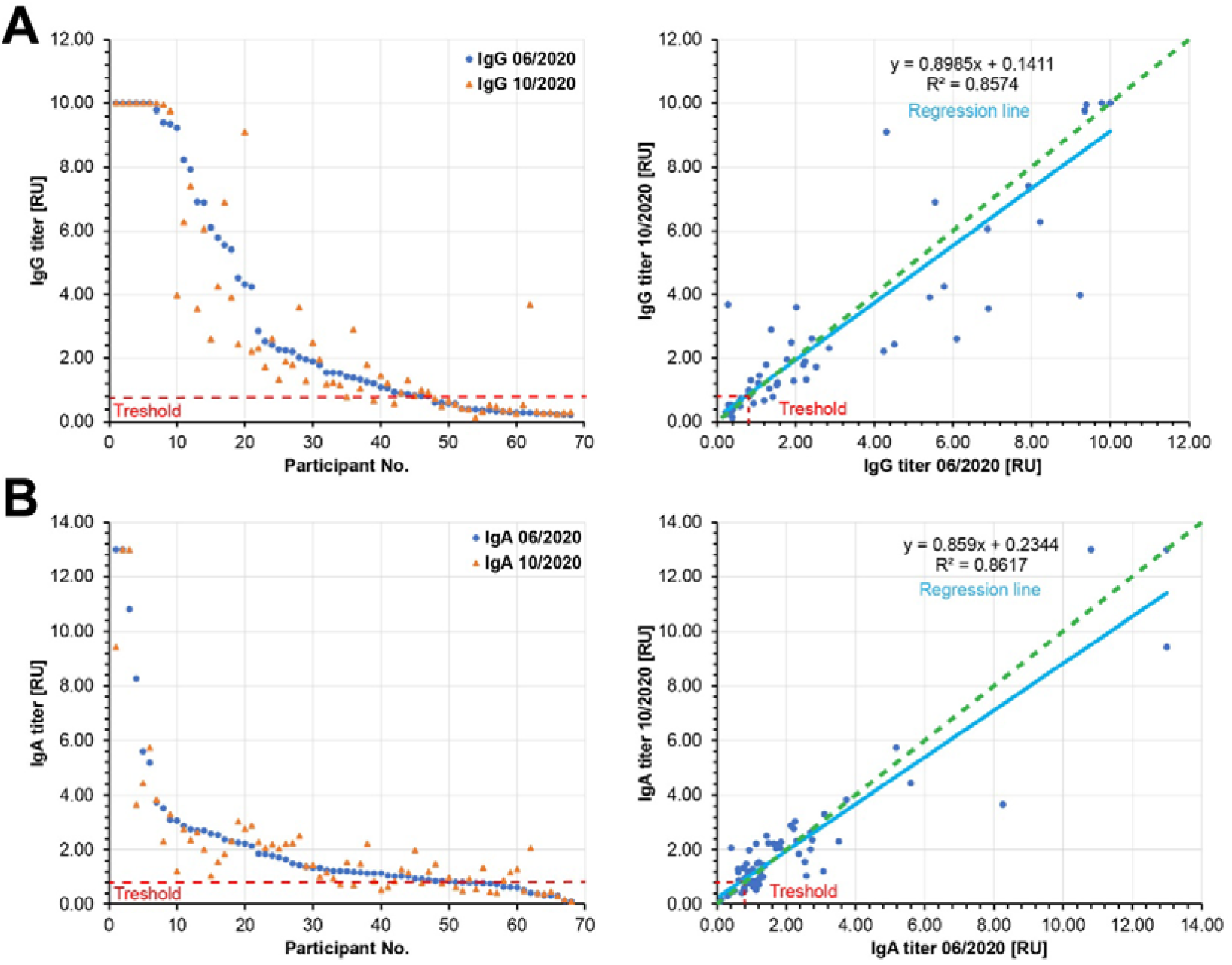
Relative SARS-CoV-2 specific IgG (A) and IgA (B) antibody titers. Left panels: The participants are ordered according to decreasing relative antibody titers of the pilot study (blue, 06/2020). The respective relatvie antibody titers of the follow-up study (10/2020) were plotted in orange. Right panels: The relative antibody titers of the follow-up study (10/2020) are plotted against the relative antibody titers of the pilot study (06/2020). The slopes of the regression lines (light blue) are below 1.0, showing a moderate waning to the relative antibody titers by 10% for IgG and by 14% for IgA. Green dashed lines: hypothetical regression lines in the case of 100% antibody persistence. Red dashed lines: Treshhold of significant antibody detection (0.8 for both IgG and IgA). Only the data of the 68 participants are shown here, whose sera were analyzed in both pilot and follow-up studies.

For how long SARS-CoV-2-specific antibody persist providing immunity is still an open debate. Several studies suggest that the immune response persists for at least several months (Baumgarth et al., 2020;Crawford et al., 2020;Isho et al., 2020;Iyer et al., 2020;Ripperger et al., 2020;Rodda et al., 2020;Wajnberg et al., 2020), whereas others propose quick waning of the SARS-CoV-2-specific antibodies in the blood serum of previously infected individuals (Long et al., 2020b;Ward et al., 2020). Our data support the idea of a prolonged immune response.

So far, studies determining antibody-based immune responses were performed with either corona antibody rapid tests, which are less sensitive, or, as in our studies, with the state of the art semiquantitative ELISA tests. Currently, ELISA methods for the quantitative assessment of SARS-CoV-2-specific IgG and IgA antibodies are emerging, which will allow a much more precise determination of antibody waning post infection. In this study samples were measured with both test systems in parallel for the comparison of the semiquantitative (see Figure 3) and quantitative analyses (data not shown and to be published later) and to set a common base for following studies.

In the light of these technological advancements and the still unclear knowledge about the stability of SARS-CoV-2-triggered antibody-based immune responses, we will continue to regularly test our cohort for SARS-CoV-2-specific IgG and IgA antibodies with both semiquantitative and quantitative ELISA, and combine these data with novel tests for SARS-CoV-2-specific T cell immunity. Waning of immune responses is to be expected, and we will test whether waning is influenced by age, sex, behavior (e.g. smoking, alcohol intake), weight, pre-existing conditions. We will also consider the role of the previous COVID-19 disease severity, as this has been proposed to influence the persistence of immunity with COVID-19 (Chen et al., 2020). So far, we could not detect any significant correlation among the persistence of antibody responses and these hallmarks, but this may change when antibody waning becomes more relevant.

## Data Availability

No supplementary to this manuscript, all original data is available on request

